# Study of Hepatitis C Virus infection among multi-transfused patients with inherited β-globin synthesis gene defect, in the eastern region of India

**DOI:** 10.1101/2020.05.06.20092049

**Authors:** Supradip Dutta, Aritra Biswas, Promisree Choudhury, Sagnik Bakshi, Prosanto Chowdhury, Maitreyee Bhattacharyya, Sharmistha Chakraborty, Shanta Dutta, Provash C Sadhukhan

## Abstract

**Background:** Post transfusion acquired HCV infection is common in high-risk group individuals such as multi-transfused β-thalassemia patients who depend on regular blood transfusions. This study was conducted to determine epidemiology and distribution of HCV in multi-transfused β-thalassemia patients, in West Bengal, India.

**Methods:** Over a span of six years blood samples were collected from HCV sero-reactive β-thalassemic patients and processed for viral RNA isolation followed by nested RT-PCR for qualitative viremia detection. HCV genotype was determined by amplifying partial HCV core gene by nested RT-PCR, DNA sequencing and using NCBI genotyping tools. Phylogenetic and phylogeographic studies were performed with online MEGA-X and BEAST 1.10.0 software respectively.

**Results:** Out of 917 multi-transfused HCV sero-reactive β-thalassemic patients, 598 (65.21%) were positive for HCV RNA while 250 (41.80%) had spontaneously cleared the virus. Female thalassemic patients and individuals belonging to ages 10-14 years had higher chances of spontaneous clearance. The most prevalent circulatory HCV genotype was 3a (78.11%) followed by 1b (12.20%). Phylogeographic analyses revealed that the 3a strains share similarity with Pakistan, Sri Lanka and Thailand whereas the 1b strains share similarity with Thailand, Vietnam, Russia and China.

**Conclusion:** The prevalence of HCV infection is very high among Indian β-thalassemic patients, necessitates a critical look into the prevailing transfusion practices and requires implementation of more rigid donor screening criteria to decrease the rate of transfusion transmitted HCV infection, especially in multi-transfused thalassemic patients. The use of more sensitive NAT based assays for HCV detection in donor blood is a compressing need of the hour.

## INTRODUCTION

HCV is an enveloped, positive sense, ∼ 9.6 kb ssRNA virus belong to Hepacivirus genus and *Flaviviridae* family that can cause chronic liver diseases, including cirrhosis of liver and hepatocellular carcinoma (HCC). HCV mainly transmits through blood, blood products and body fluids infecting an estimated 200 million people globally ^1^. After leaving the bloodstream, HCV enters the liver and replicates in hepatocytes, resulting acute or chronic liver infection. Based on viral genetic diversity, HCV has been classified into seven genotypes (1-7) and over 67 subtypes ^2^. It is extremely important to know the HCV genotype of an infected patient for genotype specific Direct Acting Antivirals (DAAs) treatment ^3^. HCV genotypes differ from each other not only in nucleotide sequences but also in geographical distribution ^4^.

β-thalassemia is an inherited autosomal recessive disorder arises from reduced or lack of synthesis of the beta globin chain of hemoglobin ^5^. β-thalassemia is prevalent all over the world; however, the frequency is higher in the Mediterranean region, the middle east, the Indian subcontinent and Southeast Asian countries ^6^. Approximately one-tenth of the world’s thalassemic patients are born in India every year of which approximately 1-3% in southern India and 3-15% in northern India are carriers ^7^. Since regular blood transfusion is the major treatment option for multi-transfused β-thalassemia patients, these patients are at a higher risk of acquiring post transfusion HCV infection, if the transfused blood was collected during the donor’s seronegative “window” period ^8^.

Genomic diversity of HCV in thalassemia patients depends on geographical localization. Different countries may have different genotype distribution^9^. In India, genotype 3 is prevalent in the north, east and west and genotype 1 in the south^10^. Although, a hospital-based study on the prevalence and genotypic distribution of HCV in thalassemia patients were conducted ^11^, there was still a lack of a more comprehensive study on the genomic diversity of HCV in β-thalassemic individuals from India. This study was undertaken to highlight the viremia, genomic diversity and evolution dynamics of HCV among multi-transfused β-thalassemia patient population in the Indian state of West Bengal from 2014-2019.

## MATERIALS AND METHODS

### Ethical Statement

Written informed consent was obtained from the patients before including them for this study. This study protocol complied with the Helsinki Declaration of 1975, amended in 2013, and was approved by the Institutional Ethical Committee, Indian Council of Medical Research-National Institute of Cholera and Enteric Diseases (ICMR-NICED), Kolkata, India.

### Study Design

A total of 917 HCV sero-reactive β -thalassemic patients were enrolled in this study. Patient’s blood samples were collected by venipuncture in clot vials followed by collection of demographic and clinical data from 10 collaborating transfusion centers (TCs). Patients with other viral co-infections were not included in this study. These 10 TCs were named as TC 1-10 and are located around eight southern districts namely, Bankura, East Burdwan, West Burdwan, East Midnapur, West Midnapur, Howrah, Hooghly and Kolkata in West Bengal, the eastern state of India. All laboratory works were done at ICMR-NICED, Kolkata. HCV sero-reactivity were rechecked by HCV ELISA (HCV Mono ELISA, BioRad). HCV RNA positive patients were classified into seven age groups (AG), 1 to 7, depending on their age namely, AG-1 (2-5 years), AG-2 (6-9 years), AG-3 (10-14 years), AG-4 (15-19 years), AG-5 (20-24 years), AG-6 (25-29 years) and AG-7 (30 years and above). Patient age ranged from 2 to 35 years with a mean (±SD) of 11.5 ± 7.5 years. Spontaneous clearance was defined as undetectable level of HCV RNA (viral load < 50 IU/ml) by quantitative Real Time PCR in two consecutive testing in 6 months apart. Patients who achieved spontaneous clearance in this study were followed up to 2 years after initial detection of infection.

### Detection of HCV RNA and genotyping

Viral RNA was isolated from all sero-reactive serum samples using QIAamp viral RNA mini kit (Qiagen, Hilden, Germany) according to manufacturer’s instructions. Briefly, viral RNA was extracted from 140μl serum, eluted in 50μl elution buffer and stored at −80°C for further use.

Detection of HCV viral RNA was done by nested RT-PCR based on 5’UTR of HCV genome as described previously ^12^. Briefly, first round one tube RT-PCR was done in 20μl total reaction volume containing 2μl isolated RNA and second round nested PCR was performed in 25μl total volume using 2μl of 1^st^ round RT-PCR product. A positive band at 256 bp in 1.5 % agarose gel stained with ethidium bromide was observed using a gel documentation system (BioRad, USA). Quantitative HCV RNA was estimated using a Qiagen quantitative real time RT-PCR (qRT-PCR) kit (QuantiFast Pathogen RT-PCR plus IC Kit). The HCV primer and probe sequences were directed against the 5’ UTR of the HCV genome^13^ using the 4^th^ WHO International Standard for HCV (NIBSC code 06/102) as standard.

For the determination of HCV genotype, nested RT-PCR amplified amplicons of partial HCV core gene (405 bp) ^14^ were gel purified and directly used for DNA sequencing analyses in an automated DNA sequencer, model 3130XL (ABI, USA) using Big Dye terminator 3.1 kit (Applied Biosystems, USA). HCV genotyping were determined by NCBI genotyping tool (https://www.ncbi.nlm.nih.gov/projects/genotyping/formpage.cgi).

### Phylogenetic analysis

Phylogenetic analysis was performed using 122 representative partial core sequences from HCV RNA positive samples and 29 reference sequences from NCBI. To investigate the evolutionary linkage among lab strains and reference strains, partial core sequences of 86 laboratory isolated 3a strains (Accession Numbers, MN635186-MN635242, MW330397-330425), twenty one 1b strains (Accession Numbers, MN617343-MN617360, MW368893-MW368895), ten 3b strains (Accession Numbers, MN605946-MN605950, MW368887-MW368891), two 3g strains (Accession numbers-MW368883,MW368885), one 3i (Accession number-MW368886) and 1a strains (Accession number-MW368892) were aligned with HCV reference strains using Molecular Evolutionary Genetics Analysis tool (MEGA-X)^15^. The evolutionary history was inferred using the Neighbor-Joining (NJ) method^16^. The evolutionary history was inferred by using the Maximum Likelihood method and Tamura-Nei model. The tree with the highest log likelihood (−3826.33) was shown. The percentage of trees in which the associated taxa clustered together is shown next to the branches. Initial tree(s) for the heuristic search were obtained automatically by applying Neighbor-Join and BioNJ algorithms to a matrix of pairwise distances estimated using the Maximum Composite Likelihood (MCL) approach, and then selecting the topology with superior log likelihood value. A discrete Gamma distribution was used to model evolutionary rate differences among sites (5 categories (+*G*, parameter = 0.4698)).

### Phylogeographic analysis

To understand the phylogeographic relationship among neighboring countries and HCV genotype distribution with evolutionary linkages in β -thalassemia population, phylogeographic trees were constructed using Bayesian Evolutionary Analysis Sampling Trees (BEAST), package 1.10.4 (http://beast.community/programs). HCV core sequences from the following countries, India, China, Pakistan, Bangladesh, Myanmar, Thailand, Japan, Vietnam, Iran, Australia were downloaded from ViPR (Virus Pathogen Resource) database (www.viprbrc.org/brc/home.spg?decorator=vipr), HCV sequence database (https://hcv.lanl.gov/content/index) and NCBI. First 300 bp (Respective of 371 nt to 671 nt of the H77-accession number NC-004102) of conserved HCV core regions were used for analysis. The General Time Reversible (GTR) model of nucleotide substitution was used with gamma + invariant sites distribution. For the clocks setting, uncorrelated relaxed clock type was used ^17^ along with the Bayesian Skyline Coalescent model. It was also ensured achievement of Effective Sampling Sizes (ESS) >200 in TRACER (http://beast.community/tracer)^18^. The result of BEAST analyses was evaluated in TRACER. Tree Annotator (https://beast.community/treeannotator) and Fig-Tree (https://beast.community/figtree) software were used to construct the final tree.

## RESULTS

### Distribution of HCV viremia in ten transfusion centers

A total of 917 HCV sero-reactive serum samples were collected over six years from multi-transfused β-globulin synthesis defective thalassemia patients at either of the 10 TCs located around ten districts of West Bengal, India as indicated in **Figure 1**. Overall, HCV RNA was detected in 65.21% (n=598) sero-reactive samples by either qualitative nested RT-PCR or quantitative real time RT-PCR. Prevalence of HCV RNA as observed in the 10 TCs were 81.25% (TC-1, n=30), 70.27% (TC-2, n=30), 70.58% (TC-3, n=28), 64.20% (TC-4, n=213), 58.51% (TC-5, n=65), 56.75% (TC-6, n=24), 82.27% (TC-7, n=75), 60.52% (TC-8, n=27), 52.56% (TC-9, n=47) and 62.33% (TC-10, n=59) respectively. The distribution of HCV viremia among the different TCs is depicted in **Table 1**.

**Table 1.**
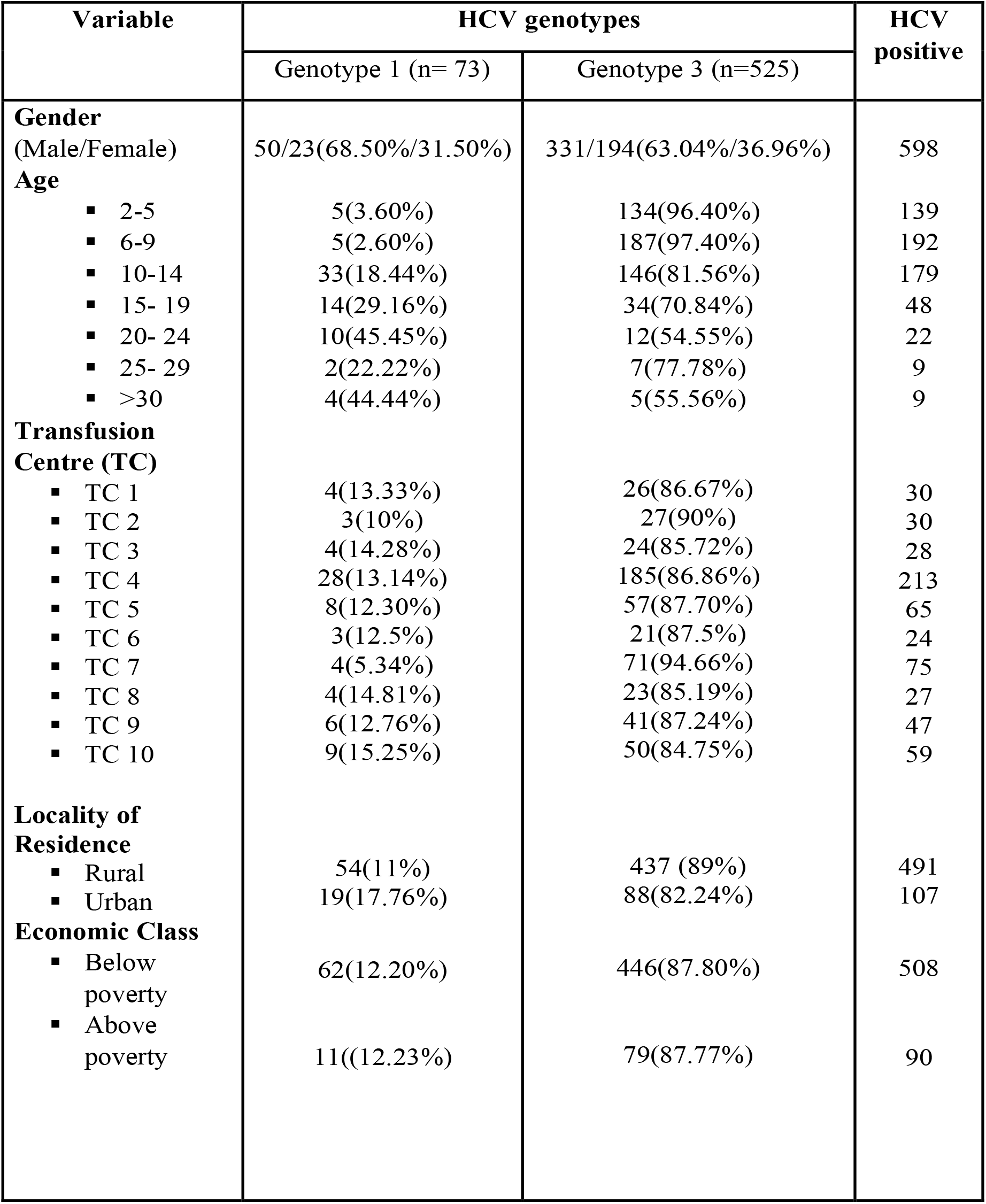

**Figure 1:**
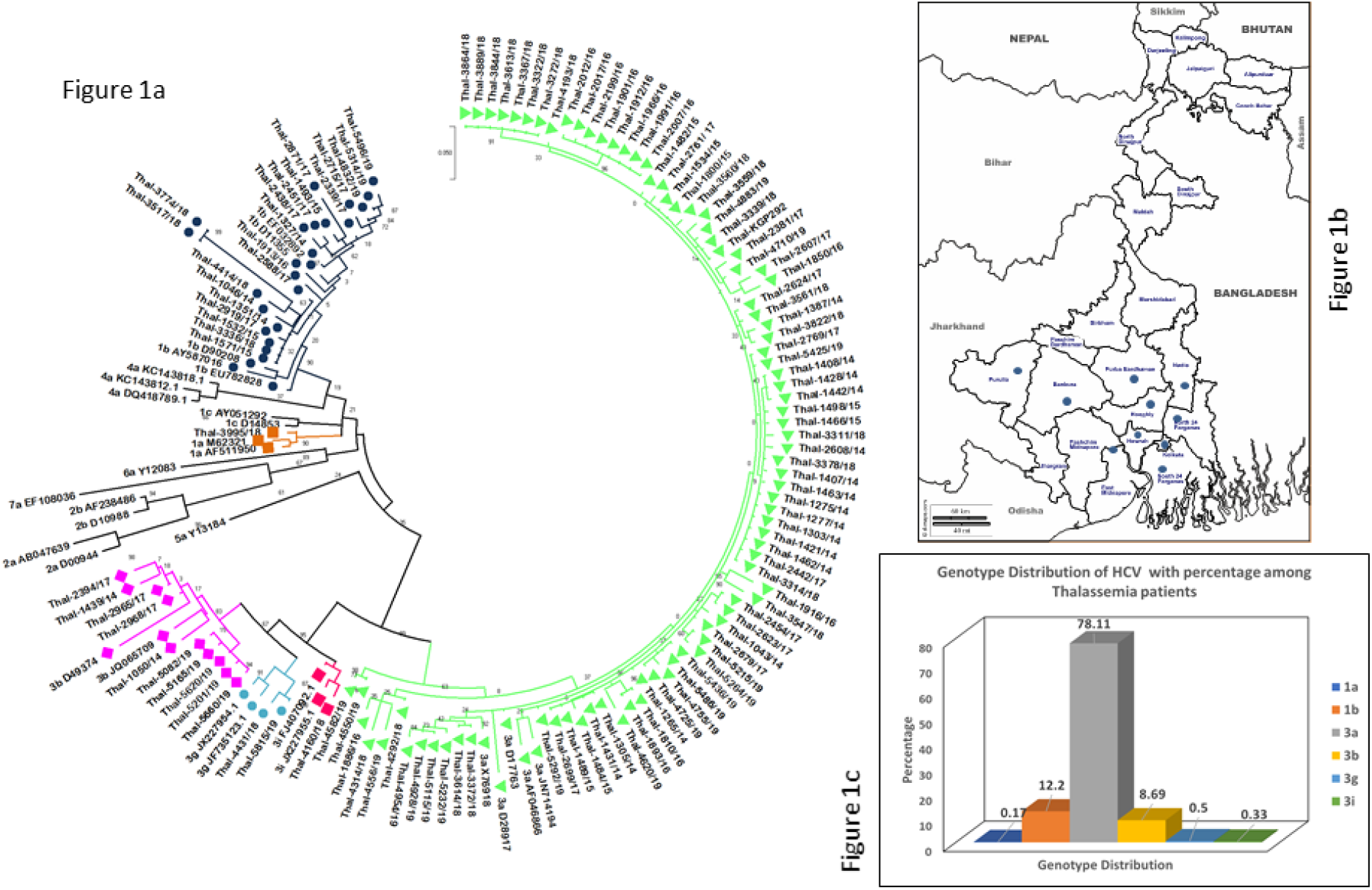
Phylogenetic tree (Fig 1a), Relative positions of Transfusion Centres in West Bengal Map (Fig 1b), Genotype distribution of HCV with percentage among thalassemia patients (Fig 1c).

### HCV genotype distribution, demographic and risk factors

598 partial core gene sequences (405 bp) were amplified and sequenced for HCV genotyping and phylogenetic analyses. Sequences were aligned with references sequences by NCBI genotyping tool and it revealed that 87.80% (n=525) patients of our study population were infected with HCV genotype 3 and 12.20% (n=73) patients were infected with HCV genotype 1. The circulating HCV subtypes that were detected in this region were: 1a, 1b, 3a, 3b, 3g, and 3i with subtype 3a being the predominant subtype (78.11%, n = 467) followed by subtype 1b (12.03%, n=72) and 3b (8.69%, n=52), while subtypes 3g, 3i and 1a were found in only 3 (0.50%), 2 (0.33%) and 1 (0.17%) patient(s) respectively. Studies on the distribution of HCV genotypes in each TC individually revealed that HCV genotype 3a was detected in all the 10 TCs (TC1-TC10) and was the most prevalent HCV strain among all these 10 centers. HCV strains 1b and 3b could be found in only seven (TC-1 to TC-7) and nine (TC-1 to TC-9) TCs respectively, whereas HCV strains 1a, 3g and 3i were detected from one (TC-6), two (TC-5, TC-9) and one (TC-7) TC(s) respectively. Population based DNA sequencing data from the partial core region of HCV genome showed no multiple HCV genotype infections in any of the β-thalassemia patients in this study population. HCV sequences reported in this study were deposited in gene bank with accession numbers as mentioned in the materials and methods section.

### Phylogeography

To examine phylogeographic similarity between India and its neighboring countries, a retrospective Bayesian dated phylogeographic analysis was performed using partial core gene sequences of HCV strains obtained from NCBI sources available online. The maximum clade credibility tree was constructed on the basis of partial core gene sequences for genotypes 3a and 1b. We found that majority of HCV genotype 3a sequences isolated in our laboratory formed three distinct clusters resulting in the Pakistan clade, Thailand clade and India/Sri Lanka clade **(Figure 2)** whereas laboratory isolated HCV genotype 1b strains were similar to isolates from Thailand, Vietnam, Russia and China **(Figure 3)**.

**Figure 2:**
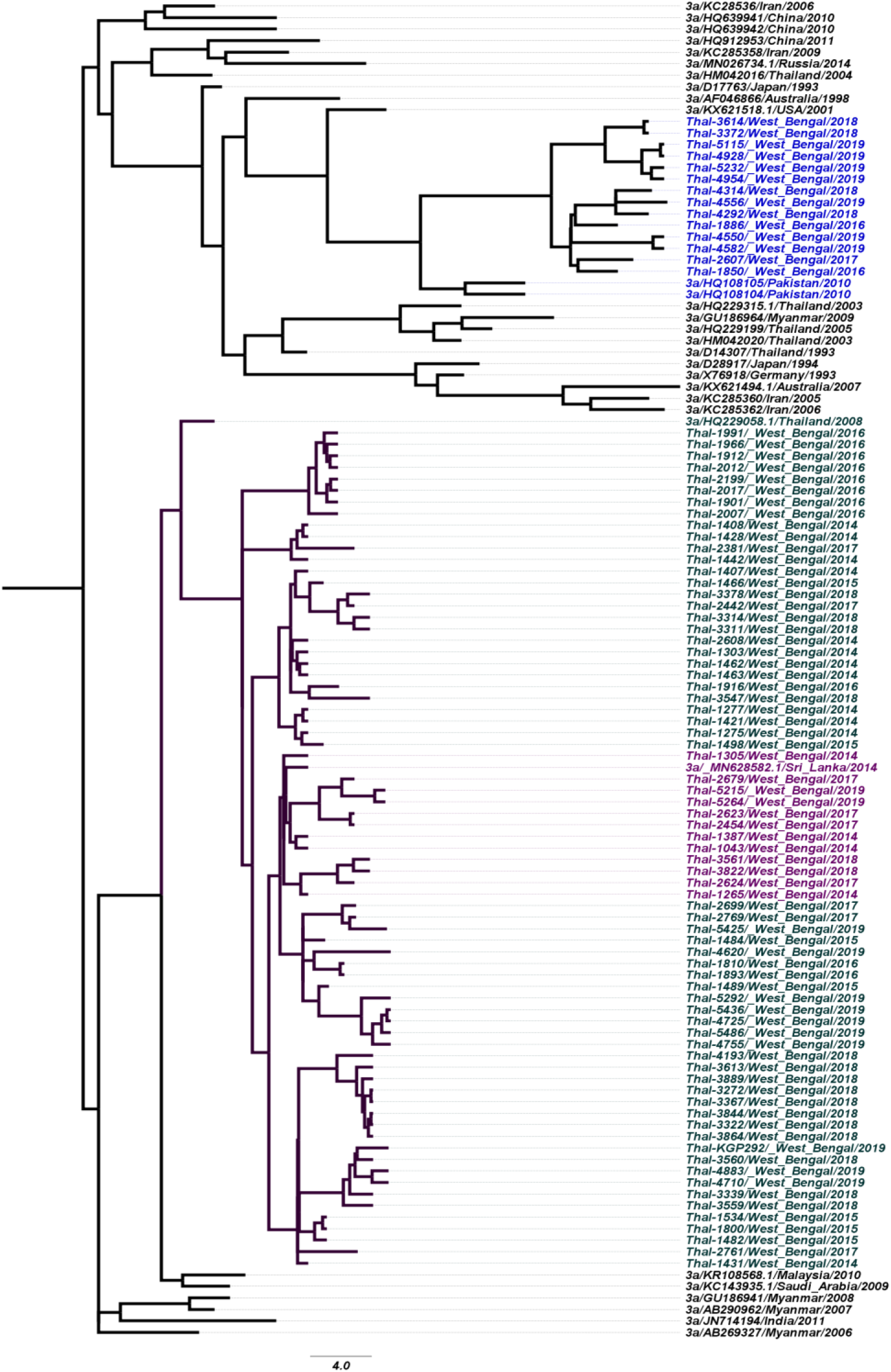
3a phylogeography

**Figure 3:**
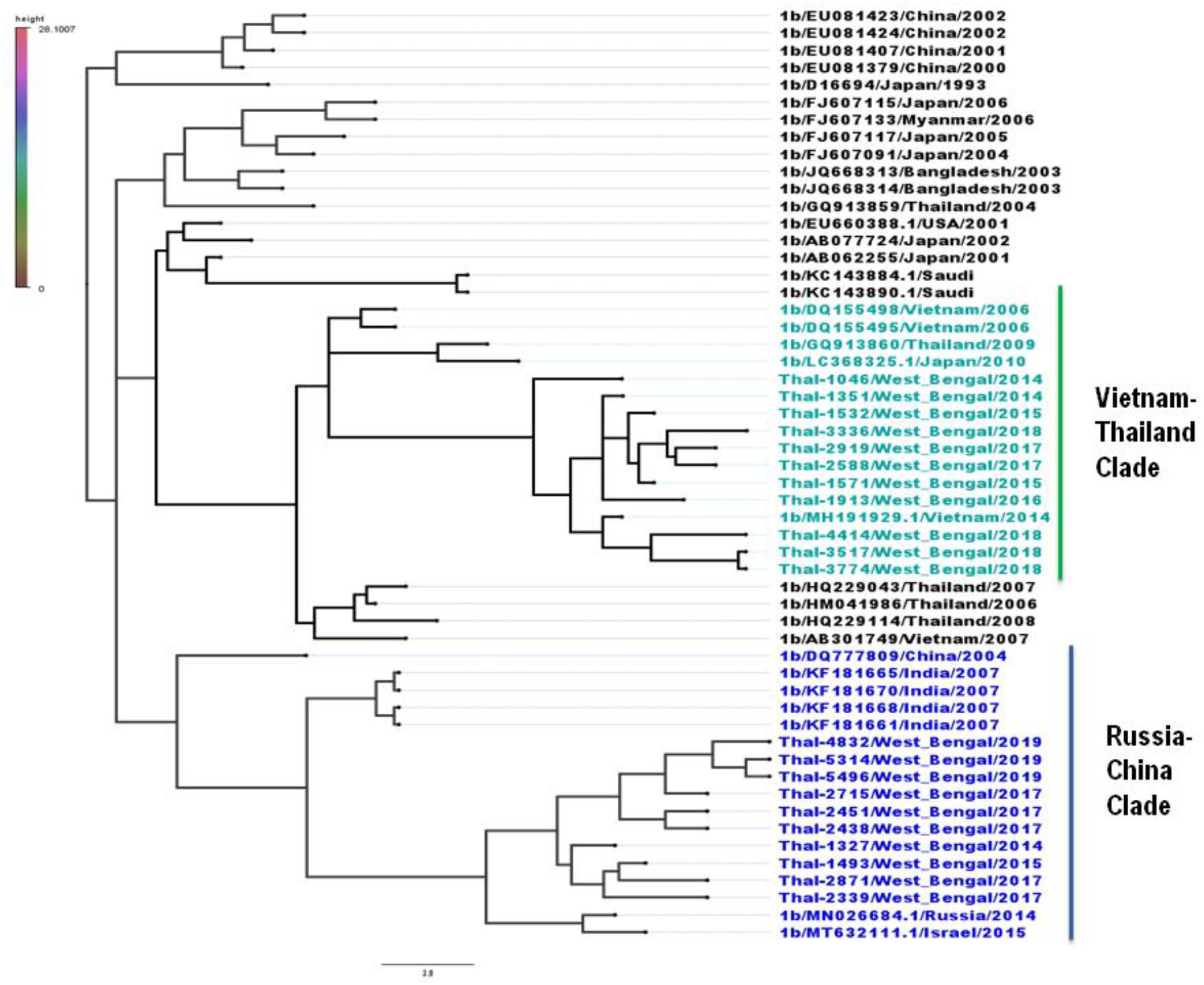
1b phylogeography

## DISCUSSION

HCV infection is one of the most common TTIs in multi-transfused β-thalassemic individuals causing serious health complications in this high-risk group population. The epidemiology and genotype distribution of HCV infection in thalassemic individuals is not very well reported from India or other countries, especially HCV genotypes and geographic distribution. Determination of HCV genotypes and subtypes is essential to know the source of HCV infection in a specified population and it is also required for clinical management, therapeutic intervention and development of an effective HCV vaccine. This study was carried out over a period of six years (January, 2014 to December, 2019) in 917 HCV sero-reactive β-thalassemic patient samples that were collected from a total of 10 transfusion centers where patients received blood from their respective districts of West Bengal on regular **(Figure 1b)**. The prevalence of active HCV infection (state of viremia) in some of these transfusion centers was as high as 82% **(Table 1)**. This data suggests that TCs may be act as hotspots for transmitting HCV infection. It can also be postulated that due to improper screening of donor’s blood during the seronegative “window” period, serological tests such as Tri-Dot assays and ELISA, which are more commonly used for donor blood screening in the blood banks are not sufficient to detect the presence of active HCV infection in donor blood. Hence, HCV infected blood is getting transfused to the thalassemic recipients. Therefore, maintenance of strict transfusion protocols using more sensitive NAT based RT-PCR screening methods are essential to check donor’s blood for probable infections before it is used for transfusion to prevent the transmission of HCV in this high-risk group population.

Data from this study revealed that β-thalassemic patients within the age group of 6-9 years had a higher prevalence of HCV viremia **(Table 1)** which is very rarely reported and age group of 10-14 years had a higher percentage of HCV clearance **(Table 2)**. We also found that patients with lower age (6-14 years) had higher HCV clearance as compared to patients with higher age (20-29 years) suggesting that low age thalassemic patients have a higher probability of clearing the infection than older patients. This could be because of the increase in serum ferritin levels in older thalassemic patients due to repeated blood transfusions that facilitates HCV replication^19^ and leads to faster progression to liver cirrhosis and HCC^20^.

**Table 2.**

|  | Age<br>2yr-5yr | Age<br>6yr-9yr | Age<br>10yr-14yr | Age<br>15yr-19yr | Age<br>20yr-24yr | Total number<br>of<br>Spontaneous<br>Clearance |
| --- | --- | --- | --- | --- | --- | --- |
| Male<br>(total no. of<br>follow-ups,<br>n= 309 out<br>of 381<br>patients) | 30<br>(22.06%) | 45<br>(33.09%) | 52<br>(38.24%) | 6<br>(4.41%) | 3<br>(2.20%) | 136<br>(44%) |
| Female<br>(total no. of<br>follow-ups,<br>n= 176 out<br>of 217<br>patients) | 25<br>(21.93%) | 39<br>(34.22%) | 43<br>(37.72%) | 5<br>(4.38%) | 2<br>(1.75%) | 114<br>(65%) |

It has been previously reported that women can clear acute HCV infection at a higher rate than men^21^. Data from this study also provides evidence for the first time that in HCV infected multi-transfused β-thalassemic Indian patients, thalassemic females were more likely to spontaneously clear the virus than thalassemic males **(Table 2)**. This difference in HCV clearance rate based on gender is still an unresolved issue but there are a few plausible explanations. In most viral infections, basal immune responses are higher in females than in males and low doses of estrogen have been suggested to modulate antiviral innate and adaptive immune responses^22^. Secondly, women have a slower rate of liver disease progression including liver cirrhosis and HCC^23^, thus it is not surprising that the burden of HCV disease complications and the rate of infection clearance is lower in thalassemic females as compared to thalassemic males. In addition to these factors, several other yet to be determined like, genetic and immunologic factors may play a key role in favoring HCV clearance in the thalassemic females.

In this study, six HCV subtypes were noticed and they were 1a, 1b, 3a, 3b, 3g and 3i **(Figure 1a, 1c)** of which genotypes 3g and 3i have not been previously reported from this region and this is the first time that these two HCV subtypes were found circulating in β-thalassemic patients of West Bengal. Another interesting finding is the higher predominance of HCV genotype 3 (specially subtype 3a) among β -thalassemic patients irrespective of the fact that the transfusion centers from where these patients received blood transfusions were all located in different districts. We had also observed similar pattern of HCV genotype 3 (subtype 3a) predominance among β-thalassemic patients in Iran^24^. It was also found that HCV subtype 3a mainly distributed through central, middle-east, and south-east Asian countries^25-30^when almost all these countries were situated on ‘Global Thalassemia belt’^31^. In South American sub-continent thalassemia cases can be found in Brazil^32^ and with more surprising observation that HCV genotype 3a was also present over there in a relatively high rate^33^. Although the predominance of HCV genotype 3 (63.85%) in the Indian population have been reported earlier^34^ but our study reports that the prevalence of HCV genotype 3 among the Indian β-thalassemic patients is even higher (87.79%). Several factors are known to influence HCV replication such as host immunity, age, hepatocellular damage, whether the infection is acute or chronic and these factors might provide a replicative advantage to one HCV genotype in a specific patient population than other genotypes^35^. Although a more detailed analyses is necessary to prove this but from our observations it can be preliminarily speculated that HCV genotype 3a has a replication priority over other HCV genotypes during the natural course of infection in the β-thalassemic Indian population. HCV genotype 3 also associated to more aggressive hepatic steatosis and liver cancer^36–38^, which may impart a serious life threat to β-thalassemia patients and further reduce their lifespan due to additional complications like iron overloading, splenomegaly, etc.

Phylogeographic analysis of HCV strains assist in monitoring the virus for molecular tracing, ancestral studies, genetic assessments, clinical and therapeutic intervention. Hence, phylogeographic analysis was performed with two most commonly found HCV genotypes among β-thalassemia patients in West Bengal, 3a and 1b to obtain information about the hierarchal relationships and genetic evolution. Data suggests that majority of the HCV 3a isolates from thalassemia patients in West Bengal have sequence similarities with HCV strains from neighboring countries like Pakistan, Sri Lanka and Thailand **(Figure 2)** whereas majority of the HCV 1b isolates of this region had sequence similarities to HCV strains of Vietnam, Thailand, Russia, and China **(Figure 3)**, suggesting that the point of origin of the circulating HCV strains 3a and 1b can be traced back to the adjacent countries that share border with India. However, we could not perform phylogeographic analyses of HCV strains 1a, 3i and 3g as we did not isolate sufficient numbers of these strains from this study population to undertake any such analytical study.

To summarize, this study provides the first multi-district, comprehensive and detail analyses of HCV viremia and genomic diversity among β-thalassemia patients in eastern part of India. Two major genotypes with 6 subtypes are circulating within this study population and the most prevalent HCV strains were 3a and 1b. This study provides the evidence of the presence of two new additional HCV strains 3i and 3g, which are not commonly reported from this region. Our study also indicated that young adults/ youth between the ages of 10-14 years and female β-thalassemic patients had a higher probability of spontaneously clearing the virus. Additionally, this study is also reported that laboratory isolated HCV strains 3a and 1b have sequence similarity with neighboring countries like, China, Pakistan, Myanmar and Bangladesh and may be those HCV strains migrated to this region.

Beta-Thalassemia patients are at very high risk for HCV infection due to repeated blood transfusion. Yet, very little studies have been reported about prevalence and genotype distribution of HCV in this neglected population which is the main focus of our study.

We have found very High HCV prevalence among beta-thalassemia patients specially in children of eastern part of India. They are also at a high risk of Cirrhosis because genotype 3 (specially 3a) is the most prevalent genotype in this population. Two new subtypes 3g and 3i also have been seen in this population which can be a future problem for genotype specific HCV Directly Acting Antivirals (DAAs) treatment and clinical management for thalassemia population in this region. Two major genotypes (3a and 1b) share similarities with neighbouring countries HCV strains.

When all World is fighting to eradicate HCV within 2030, Thalassemia population can be a major hurdle in that path, if proper clinical management and NAT based detections of donor blood are not implemented as early as possible.

## Data Availability

All data reported in this manuscript will be available on request post manuscript publication as this is an unpublished preprint.

## Acknowledgements

We would like to acknowledge all the past and present members of Dr. Sadhukhan laboratory: Kallol Saha, Rushna Firdaus, Upasana Baskey, Priya K. Verma, Raina Das, Ronita De, Sudipta Biswas and Maya Halder for all their help with this study. We would like to thank the patients and their families for their kind cooperation and participation to perform this study. We would also like to thank our collaborative institutes for their help and support. We would also like to acknowledge the financial help from Indian Council of Medical Research, New Delhi, Govt. of India; Department of Science and Technology, Govt. of West Bengal for their financial support to carry out this study.

## Notes

### Competing Interest Statement

The authors have declared no competing interest.

### Funding Statement

Funding was provided by the Indian Council of Medical Research (ICMR), India, and Department of Science and Technology (DST), Government of West Bengal, India.

### Author Declarations

Ethical Statement: Written informed consent was obtained from the patients before including them for this study. This study protocol complied with the Helsinki Declaration of 1975, amended in 2013, and was approved by the Institutional Ethical Committee, Indian Council of Medical Research-National Institute of Cholera and Enteric Diseases (ICMR-NICED), Kolkata, India.

### Summary of Updates

The manuscript has been revised to add new author along with new data from the year 2019 with completely new figures and tables.

